# Comprehensive metabolite ratio QTL mapping reveals disease relevant enzyme biology

**DOI:** 10.64898/2025.12.04.25341616

**Authors:** Sadegh Rizi, Nicolas Goss, Zoltán Kutalik, Adriaan van der Graaf

## Abstract

Metabolite ratios are valuable proxies for enzyme- and pathway activity, which are otherwise hard to capture. The genetic basis of metabolite ratios has not been systematically studied as it implies expanding the search space quadratically, leading to increased computational burden. Here, we present efficient statistical methodology to identify ratio quantitative trait loci (rQTLs), requiring only association summary statistics of metabolite measurements. In validations the methodology shows strong correlation with classically estimated ratios (median ***R*^2^ = 0.94**). Across all pairwise metabolite comparisons, 5,095 metabolite pairs contain one or more significant rQTL that exhibit far stronger associations than their constituents. The genes to which these rQTLs map are strongly enriched for enzymes (Odds ratio ranges between 4.3 and 20, depending on gene mapping strategy). Furthermore, metabolites whose ratios have rQTLs have shorter reaction distance (median=4) compared to random pairs of metabolites (median=6) (***P* = 8.0**·**10*^−^*^13^**). We identified many otherwise missed loci: 1,249 rQTLs across 53 independent loci were novel, meaning that the individual metabolites did not pass significance in the source study, highlighting that our methodology increases the number of QTLs by 21%. rQTLs often reveal enzyme activity: capturing long-chain polyunsaturated fatty acid desaturation at the *FADS2* locus (177 metabolites across 1,258 rQTLs), as well as elongation through *ELOVL2* and *ELOVL5* (23 metabolites across 27 rQTLs). Importantly, 72% of genes mapped to rQTLs were not available in pQTL studies. Furthermore, tissue-specific eQTLs confirmed that some blood rQTL associations (e.g. ones mapped to *ETFDH* in muscle tissue and *SCD* in adipose tissue) can capture processes taking place in other tissues. We further identified metabolite ratios that are likely causal biomarkers for malignant bladder neoplasms and ischaemic heart disease. Finally, we identified a novel rQTL for the cAMP-to-PFOS ratio, mapping to a missense variant in *ABCG2*, suggesting that *ABCG2* is involved in the excretion of PFOS in humans. In summary, our method is able to systematically map rQTLs which can serve as key disease biomarkers, proxy for unmeasured proteins and identify novel biology. We offer an interactive browser to explore the rQTL and metabolite ratios identified in this study: metabolite-ratio-app.athirtyone.com/

## Introduction

Large biobanks are increasingly generating large datasets of molecular measurements of DNA methylation, mRNA expression, protein abundances, and metabolite concentrations [1–5]. These molecular measurements can be used as biomarkers, help to understand human biology, and subsequently provide therapeutic avenues to treat disease.

Broad measurements of plasma metabolite concentrations in hundreds of thousands of individuals using platforms such as Nightingale [6, 7] and Metabolon [3] coupled with genotyping data have led to the identification of the genetic basis of hundreds of metabolites [2, 3]. In the past, these analyses contributed to dissecting the genetic etiology and causal genes between urate and gout [8], glucose and type 2 diabetes (T2D) [9], lipid concentrations and coronary artery disease (CAD) [2], and fructose and Crohn’s disease [10].

Metabolite concentrations are dependent on reactions and their overarching metabolic pathways [11–14]. Performing a genetic association study on raw metabolite levels can lead to excess environmental variance when the concentration of the available substrates is not accounted for. This leads to a decrease in statistical detection power [15]. Investigators often use metabolite ratios to ’normalize’ metabolite levels, and identify biomarkers that are independent of substrate availability [16–18].

Unfortunately, due to computational and storage constraints, metabolite ratios are underrepresented in quantitative trait locus (QTL) studies: determining the genetic associations of the ratio of all metabolite pairs squares the computational burden compared to single metabolite tests, each of which requires a genome-wide mixed-effect model fit.

Recent work has shown that QTL summary statistics from log-transformed measurements can be meta-analyzed in such a way that the resulting summary statistics represent those found from the ratio [19]. These findings allow for computationally efficient generation of ratio quantitative trait locus (rQTL) summary statistics, but they have not been tested in a systematic manner. In a broad application on the protein quantitative trait locus (pQTL) ratios, a 25% increase in pQTL ratio loci was observed, which has elucidated multiple examples that are relevant to drug target network analysis and cytokine binding [19].

When performing rQTL analysis, it is important to note that care must be taken to avoid non-specific ratio phenotypes [20]. Fortunately, there are methods that help to ensure that an rQTL is more informative than the associations of the contributing metabolites. For instance, through the application of the *P_gain_* statistic [15], which measures the fold decrease of an rQTL *P* value over that of the constituent metabolites. A high *P_gain_* means that the rQTL is much more significant compared to the underlying association signals of the constituent metabolites considered individually. Interestingly, rQTLs often colocalize with an enzyme located in the metabolic pathway of the metabolites under study, or even with an enzyme that directly catalyzes the reaction between the substrate product pair in the ratio [3, 21]. This is important as pQTLs can be very relevant to the pharmaceutical industry, illustrated by the industry-funded UK Biobank Pharma Proteomics Project [4], and its planned order of magnitude increase in sample size [22]. While measuring protein levels at large-scale is increasingly available, the same does not hold for enzyme activity, which is more cumbersome to measure [23].

Here, we developed a summary statistics-based rQTL mapping methodology that we applied to 1,091 metabolites from Chen *et al.* [3]. We then compared our methodology to classically calculated rQTLs, yielding solid agreement. Across the 594,595 tested metabolite ratios, 5,095 have at least one rQTL that passes our stringent significance thresholds: 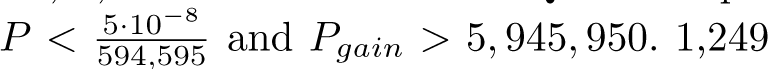 of these rQTLs were novel. The rQTLs found here recapitulate elements of metabolism, including desaturation and elongation of long polyunsaturated fatty acids as well as amino acid metabolism. Furthermore, we identify rQTLs that proxy genes beyond blood and are disease-relevant.

## Results

### Summary of the Methods

In this work, we use the summary statistics of Chen *et al.* [3] who have identified the genetics of 1,091 Metabolon-derived plasma metabolites across 8,299 individuals (**Supplementary Table 1**). Chen *et al.* have further performed a classical rQTL mapping on 299 metabolite pairs which we use as the validation set of the methodology (**Supplementary Table 1**).

Suhre [19] identified that the difference between two log-transformed metabolites is the log of their ratio and hence the difference between the summary statistic effect sizes approximates the effect size for their ratio:

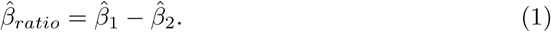

Where *β*^^^_1_ and *β*^^^_2_ are the marginal association summary statistics of the two metabolites under investigation. The standard error of this difference statistic is:

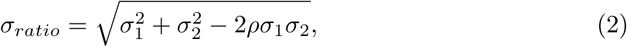

where *σ_i_* is the standard error of the respective estimate. The correlation term *ρ* is generally not known for each combination of ratios, so we built an approach to optimize *ρ* so that the genomic inflation factor for the ratio statistics equals to one. Another appropriate way of estimating *ρ* would be to use the LD score regression intercept, all the while the *ρ* estimates were highly concordant with our genomic inflation factor estimation (Pearson *ρ* = 0.992) [24] (**Supplementary Table 2**) (**Methods**). This approach is computationally beneficial, as none of the equations requires individual-level data for any of the underlying metabolites (**Methods**), which enables its application to hundreds of thousands of ratios.

### Validation of Summary Statistic Ratio Methodology

We validated our metabolite ratio methodology on 299 classically calculated metabolite ratio summary statistics between two metabolites as reported by Chen *et al.* [3]. Here, we compared the effect sizes and *Z* statistics between the originally reported ratio summary statistics and the ones derived from our methodology to ensure that our approach is reliable and controls type I error (**Methods**). As a first example, we validated the ratio between alpha-ketobutyrate and 3-methyl-2-oxovalerate. The derived effect sizes (when both the original ratio and the derived methodology are *P <* 0.05) (*β*) are highly correlated with the true ones (Pearson *R*^2^ = 0.97) (**Figure 1a**) (**Supplementary Table 3**), while the resulting 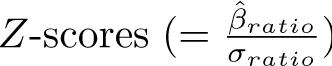, have similar concordance (Pearson *R*^2^ = 0.97) (**Figure 1b**) (**Supplementary Table 3**).

**Fig. 1.**
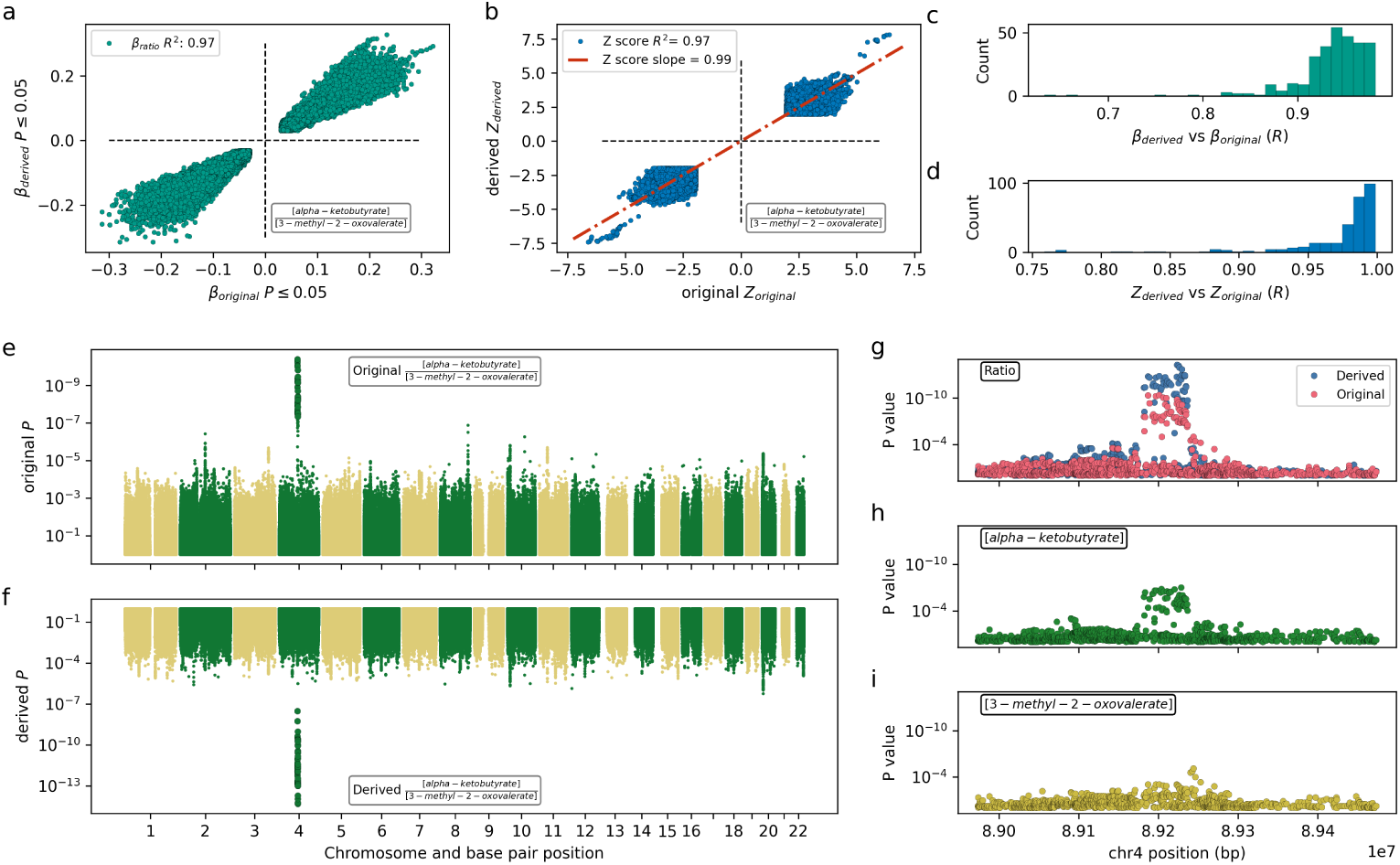
Validation of the metabolite ratio method. a) The effect size comparison of a classically calculated *β_original_* ratio compared to the derived ratio QTL *β_derived_* (this method), when the *P ≤* 0.05 for both methods. Shown here is the ratio of alpha-ketobutyrate and 3-methyl-2-oxovalerate b) The Z-score comparison of a classically calculated ratio *Z_original_* compared to our derived method *Z_ratio_*, when the *P ≤* 0.05 for both methods. c) Histograms of the Pearson correlations between the effect sizes of classically calculated ratios with derived ratios. d) Histogram of the squared Pearson correlations between *Z_original_* versus *Z_derived_* for all 299 possible ratios from which we can validate. e) Manhattan plot of original ratio between alpha-ketobutyrate and 3-methyl-2-oxovalerate. f) Manhattan plot of the derived ratio of alpha-ketobutyrate and 3-methyl-2-oxovalerate. g) Locus zoom plot of the original ratio of alpha-ketobutyrate and 3-methyl-2-oxovalerate (in red), and the derived ratio (in blue). h) The individual locus plot associations of alpha-ketobutyrate. i) The individual locus plot associations of 3-methyl-2-oxovalerate.

We extended these correlation coefficients to all 299 possible validation datasets where gold standard ratio summary statistics were available. Overall, we see that the *β* effect sizes are correlated with a Pearson *R*^2^ that ranges between 0.63 and 0.98 (**Figure 1c**) (**Supplementary Table 3**) with a median of 0.94. For Z-scores this correlation range was [0.75 *−* 0.99] with a median of 0.99 (**Figure 1d**)

Although the correlation between *β* and *Z* scores may be strong, we tested that there are no spurious loci that pass our P value threshold. E.g., for the alpha-ketobutyrate to 3-methyl-2-oxovalerate ratio, we identify the same locus as the original analysis (**Figure 1e-f**). To further ensure that the rQTLs that we identify using our derived method are similar, we compared all the loci that passed the Chen *et al.* [3]

Bonferroni multiple testing correction P values *P <* 4.58 *·* 10*^−^*^11^, and find that we re-identify *155* of the *196* (79 %) rQTL loci (**Supplementary Table 4**). Reassuringly, we only identified 12 loci that were not reported by Chen *et al.*, all of which were at least genome wide significant (*P <* 5 *·* 10*^−^*^8^) in the original study (**Supplementary Table 4**) (**Methods**). This leads us to believe that the derived method is conservative in identifying associated loci, and provides results similar to those obtained explicitly from GWAS of ratios.

### Identification of robust rQTL across all comparisons of 1,091 metabolites

Considering the strong correlation between the original and derived ratios, and its robust control of type I error, we performed pairwise rQTL mapping between all possible pairs of 1,091 metabolites, using our summary statistic-derived analysis. To reduce the computational burden, once we tested an A/B ratio, we did not repeat it for B/A since, by definition, its test statistic can simply be obtained by a sign flip. Analyzing the lower triangle of the pairwise comparison matrix, leaves 594,595 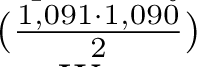 ratio comparisons.

We set conservative metabolite ratio thresholds to ensure that we do not identify spurious ratio associations through two steps: First, we only consider a SNP to be an rQTL when its association *P*-value is below the Bonferroni corrected 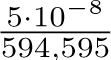 threshold. Second, we require the SNP to have an rQTL *P* value that is 10 *·* 594, 595 times smaller than the marginal association *P* value of both constituent metabolites. That is, 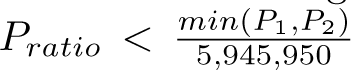[15]. This *P_gain_* statistic ensures that an rQTL identified is specific to the ratio and not simply driven by an association with one of the constituent metabolites.

Across all 594,595 tested metabolite ratios, we identified 5,095 that have at least one rQTL that satisfies both of the above-mentioned criteria (**Supplementary Table 5**). In some cases, the ratio of two metabolites has multiple associated loci, resulting in 5,954 metabolite ratio associations. Across rQTLs, we assigned 1,249 (21%) to be novel, i.e. these rQTLs contain a metabolite pair, where none of the mQTLs passed the Bonferroni significance threshold of Chen *et al.*, indicating that this locus would not have been found by simple mQTL association (**Methods**) [3]. We provide an interactive browser for all the rQTLs found in this study at metabolite-ratio-app.athirtyone.com/

These rQTLs are often located in the same region, and we have, therefore, clustered rQTLs when they are within 0.1 *R*^2^ linkage disequilibrium (LD) with one another, resulting in 210 independently associated rQTL clusters (**Figure 2a**) (**Supplementary Table 5**) (**Methods**). These exhibit variable levels of pleiotropy: 67 clustered loci are associated only with a single ratio (**Figure 2b**), while three clusters have more than 100 metabolites involved in at least one associated ratio (**Figure 2b**). Fifty-three out of the 210 (25%) rQTL clusters do not contain metabolites that passed significance in the original study, making these loci previously unreported from this data (**Methods**) (**Figure 2a**).

**Fig. 2.**
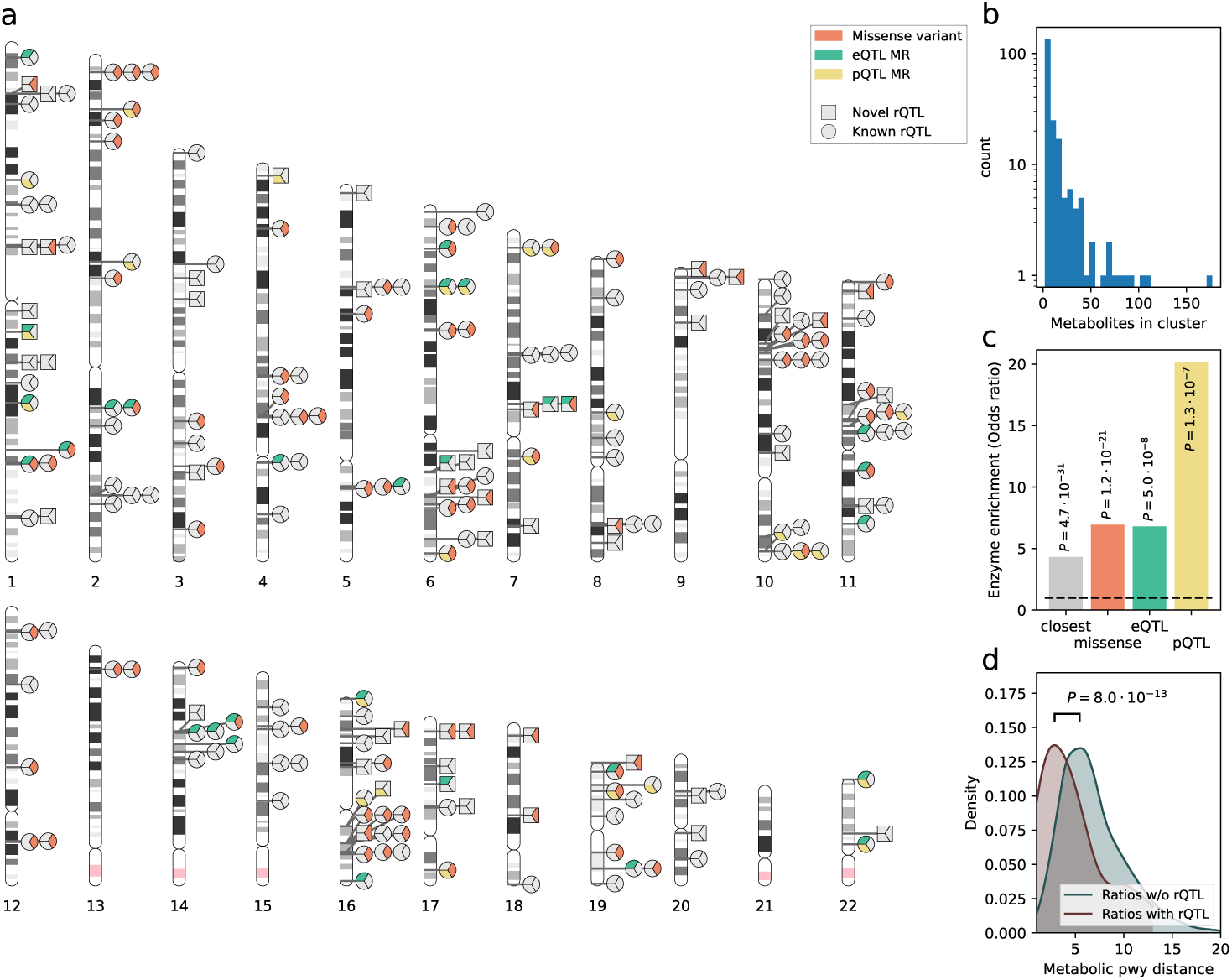
Overview of rQTL identified in this study, the enrichment for enzymes and reaction graph distance. a) rQTL ideogram showing all rQTL clusters (rQTLs that are mapped within *R*^2^ *≥* 0.1 LD), indicating their novelty compared to the constituent metabolite ratios (shape of the indicator) and if there is evidence that they potentially tag a missense variant (orange mark), an eQTL (green mark) or a pQTL (yellow mark). b) The number of metabolite ratios in each cluster (y-axis is log scaled) c) The enrichment for enzymes of each gene mapping (missense, eQTL, pQTL) method using a Fisher exact test (Methods). d) The reaction distance of metabolite pairs, depending on if their ratio has an rQTL or not.

To identify and understand the underlying genes for each rQTL cluster, we performed four gene mapping approaches: i) The closest gene mapping. ii) missense variant mapping, iii) expression QTL (eQTL) MR and iv) pQTL MR (**Figure 2a**) (**Methods**) (**Supplementary Table 6**). In missense mapping, we matched missense variants from the Ensembl variant effect predictor that were in LD *R*^2^ *≥* 0.8 of a mapped rQTL [25]. For the eQTL MR analysis, we analyzed genes from Võsa *et al.* [26] within 250 kB of the rQTL and performed four *cis*-MR and 2 colocalization methods, marking significance when the *cis*-MR method MR-link-2 identifies causality accounting for the number of testable genes at the locus 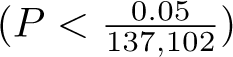 based on the conservative nature of the MR method [27] (**Methods**) (**Supplementary Table 7**). In the pQTL MR analysis, we used pQTLs from Sun *et al.* [4] using the same criteria as for the eQTL MR analysis, but as there are less proteins and thus less comparisons, the multiple testing threshold is more lenient 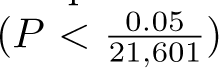 (**Methods**) (**Figure 2a**) (**Supplementary Table 8**).

Mapping rQTLs to genes allows the estimation of the likely causal gene underlying the rQTL signal. We matched these mapped genes with the Uniprot resource [28] (**Methods**) and to determine if the mapped genes are enriched for enzymes, we compared the prevalence of enzymes in the mapped genes with those among all possible genes (all genes for closest-, and missense genes, and all testable genes for eQTL and pQTL). A Fisher exact test established that they are enriched for being enzymes (Odds ratio range: 4.3-20, max *P* = 1.3 *·* 10*^−^*^7^, depending on the applied mapping method) (**Figure 2c**). Interestingly, 17 / 20 (85%) unique proteins from pQTL that MR-link-2 identified as significant are enzymes (**Supplementary Table 9**) (**Methods**).

To further ensure that the ratios having rQTLs involve metabolites that are linked via metabolic pathways, we determined the metabolic pathway distance of metabolite ratios that have an rQTL. We contrasted this reaction distance to a control set of metabolite pairs that do not have an rQTL together. The control set only contained metabolites that were otherwise present in the metabolite ratios that contain an rQTL to ensure similarity. Distances were computed using pathways from Meta-cyc, KEGG and WikiPathways based on previous work [11, 12, 14, 27] (**Methods**) (**Supplementary Table 10**). Here, we find that the median reaction distance between metabolites whose ratios have an rQTL (median=4) and ratios that do not have an rQTL (median=6) is significantly different (Kruskal-Wallis *P* = 8.0 *·* 10*^−^*^13^) (**Figure 2d**).

### rQTL in *FADS* and *ELOVL* recapitulate long polyunsaturated fatty acid metabolism

We were intrigued by the large number of rQTLs that could be found in some rQTL clusters (**Figure 2a-b**). The largest rQTL cluster was found at 11q12-q13.2, in this cluster, 177 metabolites have 1,258 individual rQTLs with one another, mapping to the *FADS1* /*FADS2* locus (**Figure 3a**). These genes encode enzymes that catalyze fatty acid desaturation (introducing double bonds to the carbon chains of fatty acids) and, hence, are involved in the biosynthesis of (poly-)unsaturated fatty acids. Common *FADS1* /*FADS2* substrates include linoleate, and palmitoate [29, 30]. Across 134 HMDB-mappable metabolites in the *FADS* cluster, 118 (88%) belong to the ’Lipids and lipid-like molecules’ superclass according to HMDB, in line with the lipid activity of the FADS enzymes [29, 31] (**Methods**).

**Fig. 3.**
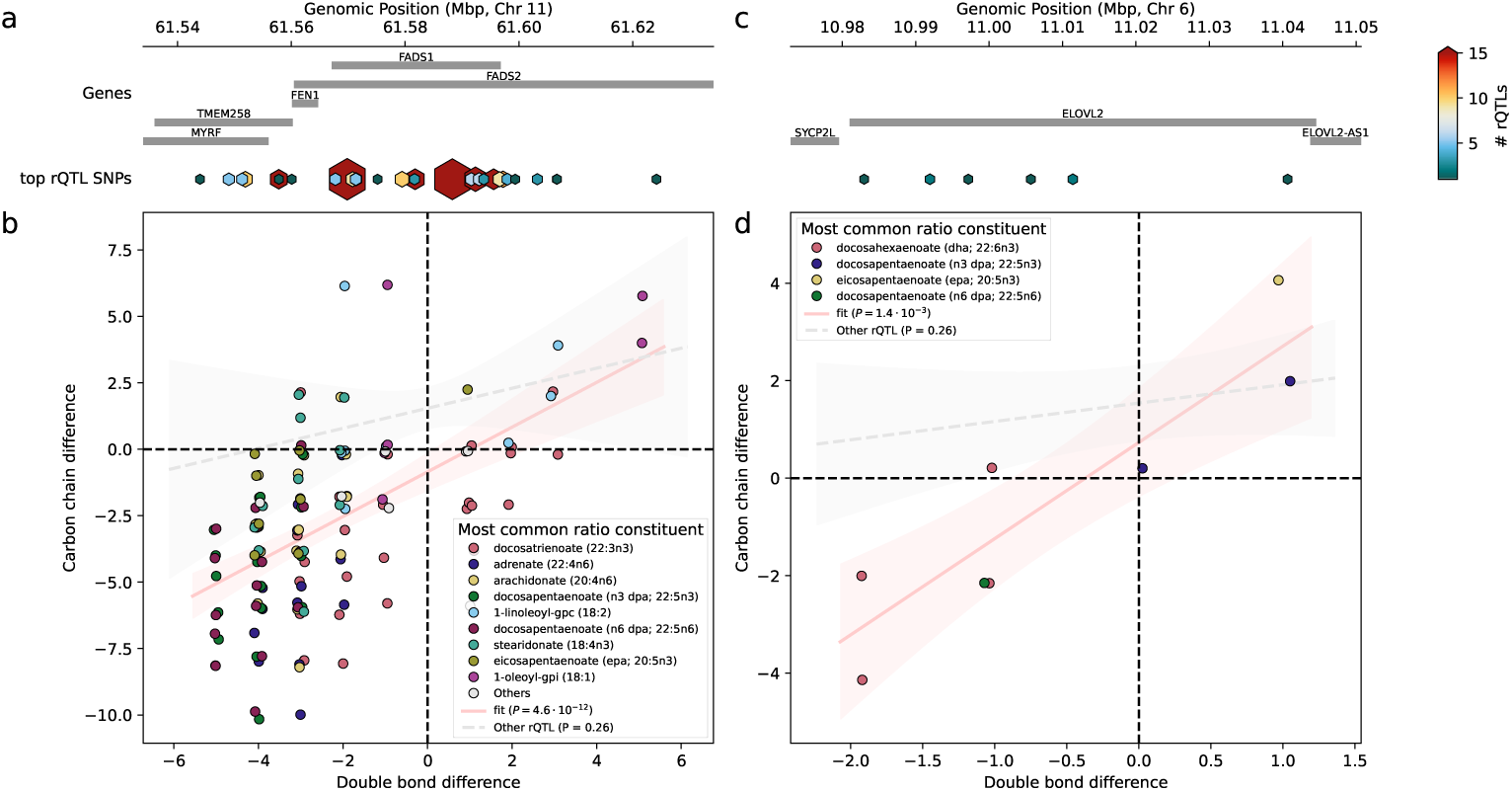
a) The *FADS* gene cluster on 11q12-q13.2, containing 132 individual ratio quantitative trait loci (rQTLs) with unambiguously saturated single chain fatty acids within linkage disequilibrium *R*^2^ *≤* 0.1. The location of the top SNPs are represented by the hexagons, colored and sized by the amount of rQTL associations. b) Unambiguous, single chain fatty acids with an rQTL in the *FADS* gene cluster. Comparing the number of double bond differences using the most commonly occuring fatty acid as the reference (colors of the points), to ensure that points are not obscured, note that points are jittered. Colored by the most commonly occurring metabolite in the rQTL comparison, and denoted ’Other’ when the relative occurrence rank is higher than 10. Regression lines denote linear regression and 95% confidence interval between the null dataset (grey) (based on monoacylic ratios with an rQTL outside of *FADS* and *ELOVL2* clusters), and the data shown on the plot. c) The *ELOVL2* locus on 6p24.2, containing 8 rQTL with unambiguously saturated single chain fatty acids within linkage disequilibrium *R*^2^ *≤* 0.1 of one another. The location of the top SNPs are represented by the hexagons, colored and sized by the amount of rQTL associations. d) Unambiguous, single chain fatty acids with a rQTL in the *ELOVL2* gene cluster, colored by the most commonly occuring fatty acid as the reference. Note that points are jittered. Regression lines denote linear regression and 95% confidence interval between the null dataset (grey) (based on monoacylic ratios with an rQTL outside of *FADS* and *ELOVL2* clusters), and the data shown on the plot (red).

We set out to identify *FADS* dependent saturation differences between metabolite pairs in the *FADS* cluster. For this, we limited the analysis to a subset of 139 *FADS* locus rQTLs where the metabolite pairs both have a single fatty acid chain and have an unambiguous number of carbons and saturations. This allowed for the comparison between metabolites in a pair for differences in the number of double bonds and the length of the carbon chain (**Methods**) (**Supplementary Table 11**) (**Figure 3b**). Interestingly, we do not identify metabolite pairs with an rQTL in the *FADS* gene cluster where the number of double bonds of the numerator and denominator single chain fatty acids is the same, aligning with the desaturation function of the FADS enzyme (**Figure 3b**). When comparing this to all metabolite pairs outside the *FADS* cluster, we identify a significant depletion of these zero double bond differences (Fisher exact test *P* = 2 *·* 10*^−^*^8^). Besides this, we find a strong and positive correlation (linear regression slope=0.84, *P* = 4.6 *·* 10*^−^*^12^) between the number of double bond difference, and the carbon chain length difference of ratios within the rQTL cluster (**Figure 3b**). This follows our expectation as the FADS enzyme works in co-regulation with fatty acid elongation enzymes to produce (poly-)unsaturated fatty acids [29, 32].

As we found a positive correlation between the number of carbons and double bonds in the fatty acids across the *FADS* locus, we investigated if we could also identify the activity of elongation enzymes. The elongation of long poly unsaturated fatty acids is specifically done by *ELOVL2* and *ELOVL5*, which are very-long-chain fatty acid elongase (ELOVL) enzymes. Our derived ratio methodology identified 24 rQTL near *ELOVL2* (**Figure 3c**) and 3 rQTL near *ELOVL5*, and no other rQTL close to *ELOVL* genes. Following literature evidence that *ELOVL2* and *ELOVL5* are the only known enzymes involved in the elongation of polyunsaturated fatty acids [33] (**Supplementary Table 11**). Some metabolite ratios have an rQTL both in the *FADS* and *ELOVL* clusters: Five out of 24 metabolite ratio pairs with rQTL in *ELOVL2* also have an exclusive rQTL in the *FADS* gene cluster, further suggesting a shared regulatory function (**Supplementary Table 11**). We selected 8 metabolite pairs containing a single carbon chain with unambiguous double bonds with rQTLs in the *ELOVL2* locus (comprising 7 unique metabolites) that do not contain ambiguous double bonds. Here, we identify an even stronger slope relationship between number of double bonds and number of carbon atoms in the fatty acid chain than in the *FADS* gene cluster (linear regression slope = 2.0, *P* = 0.0014) (**Figure 3d**). ELOVL2 elongates the carbon chain by two carbons, which is in line with the metabolite ratios with an rQTL in *ELOVL2* involving monoacylic metabolites, as they always have an even carbon chain difference. Although this observation does not pass marginal statistical significance, when compared to the single carbon chain, unambiguous double bond metabolite ratios with an rQTL outside of *ELOVL* and *FADS* (Fisher exact test: P=0.096).

Taken together, these results in the *FADS* and the *ELOVL2* rQTL clusters show that rQTLs are able to identify the broad strokes of the canonical long fatty acid elongation and desaturation pathways.

### rQTL around asparaginase and glutaminase are substrate specific

Somewhat impressively, we found that rQTL can be very specific to relevant metabolite pairs, even when they are in the same rQTL cluster: For instance, we find 7 rQTL in a small cluster close to the Asparaginase (*ASPG*) gene (**Supplementary Table 5**). Two metabolites are individually significant in the *ASPG* locus: Genetic polymorphisms for asparagine (rs12587599) and n-acetylasparagine (rs34362765) have an mQTL that is independent (LD: *R*^2^ = 0.26). When analyzing the significant rQTLs in *ASPG*, we identified one rQTL (rs8012505) for the asparagine-to-threonine ratio (*P* = 1 *·* 10*^−^*^194^), which is an *ASPG* missense variant (**Figure 4a**). Interestingly, one of its variants in LD (rs12587599, *R*^2^ = 0.96) is an rQTL for the n-acetylasparagine to n-acetylcitrulline ratio (**Figure 4b**) (**Supplementary Table 5**). The specificity of these rQTLs is illustrated by the fact that rQTLs at this locus only include the ratio of n-acetylasparagine with uniquely n-acetyl containing amino acids or similar small molecules. In summary, this locus regulates ratios of two types (**Figure 4c**).

**Fig. 4.**
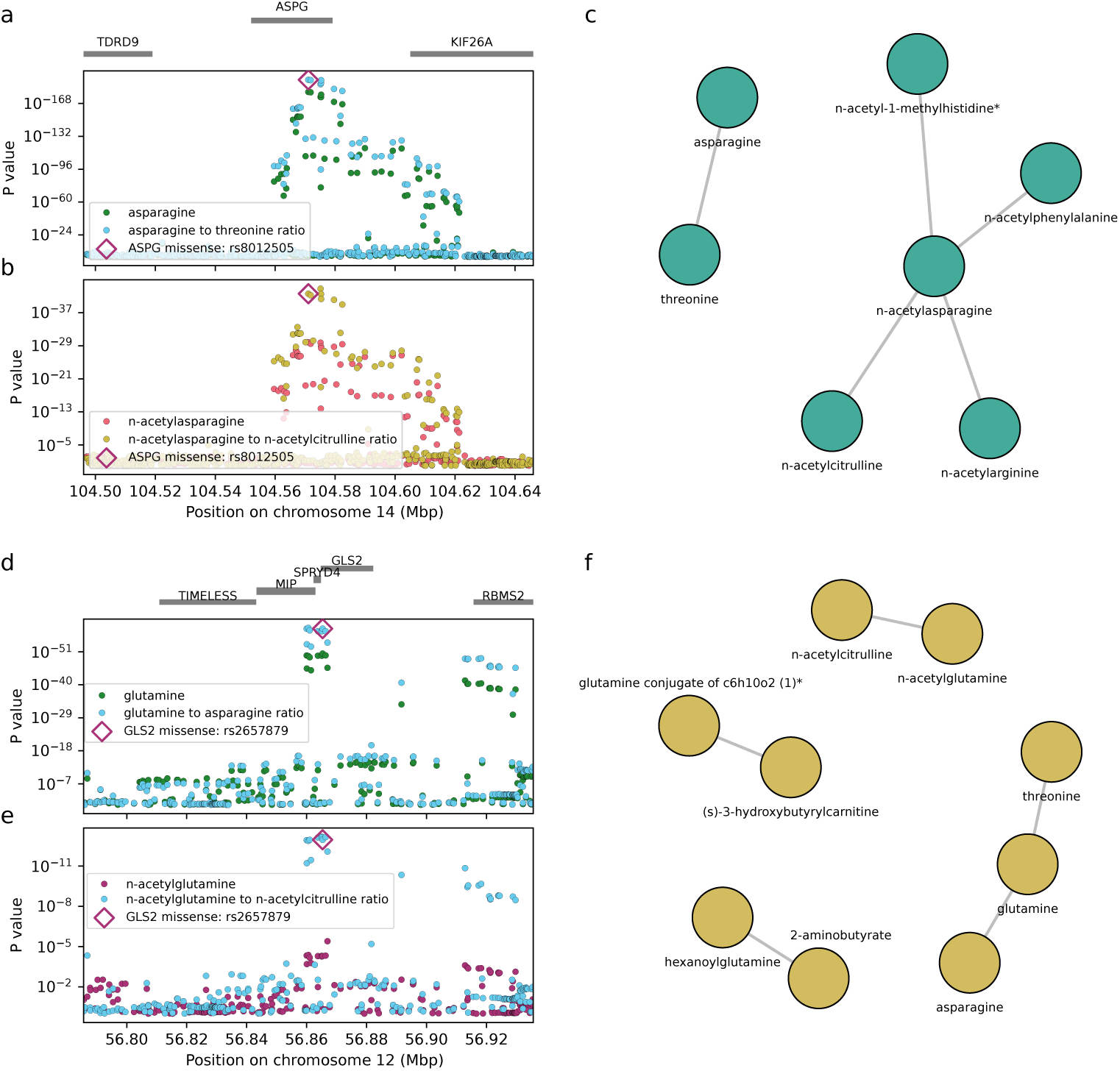
a) The locus plots of the mQTL in asparagine and the rQTL between asparagine and threonine. With the *ASPG* missense variant highlighted b) The locus plots of the mQTL in n-acetylasparagine and the rQTL between n-acetylasparagine and n-acetylcitruline. *ASPG* misssense variant highlighted c) The cluster of rQTL that are within linkage disequilibrium *R*^2^ *>* 0.1 together. d) The locus plots of glutamine- and glutamine-to-threonine ratio with the *GLS2* misssense variant highlighted. e) The locus plots of n-acetylglutamine and the n-acetylglutamine to n-acetylcitruline ratio with the *GLS2* misssense variant highlighted. f) The cluster of rQTLs around the *GLS2* that are within linkage disequilibrium *R*^2^ *>* 0.1 together.

This pattern is not only limited to the *ASPG* rQTL cluster. We find a similar pattern in the Glutaminase 2 (*GLS2*) gene cluster (**Figure 4d-f**). In this rQTL cluster on chromosome 12 our methodology identified 5 rQTLs involving 5 metabolites (**Supplementary Table 5**). Two rQTL are in high LD with an already discovered mQTL for glutamine that perfectly maps to a *GLS2* missense variant (rs2657879, *P* = 4.0 *·* 10*^−^*^51^), which is reassuring as glutamine is the substrate of Glutaminase 2. In our ratio analysis, we identify rQTLs for the glutamine to threonine ratio (rs2657879, *P* = 1.9 *·* 10*^−^*^66^) and for the asparagine to glutamine ratio (rs2933243, *P* = 1.1 *·* 10*^−^*^59^). These SNPs are in LD *R*^2^ *>* 0.98 with the missense variant in *GLS2* (**Supplementary Table 5, 6**). The 3 remaining rQTLs in the cluster are novel and the ratios associated to the SNPs do not comprise glutamine, however, each ratio contains one metabolite with a glutamine component. The ratio between hexanoylglutamine and 2-aminobutyrate has an rQTL at rs7302925 (*P* = 1.0 *·* 10*^−^*^14^), the ratio between n-acetylglutamine to n-acetylcitrulline has an rQTL at rs1043011 (*P* = 6.6 *·* 10*^−^*^14^) and the ratio between (s)-3-hydroxybutyrylcarnitine to glutamine conjugate of c6h10o2 (1)* has an rQTL at rs2631372 (*P* = 7.9 *·* 10*^−^*^23^) (**Supplementary Table 5, 6**). All three of these SNPs are in *R*^2^ *≥* 0.91 LD of the *GLS2* missense variant, however strikingly, there is no link to be made between the separate glutamine containing metabolites: The metabolite ratios are not connected to each other in a larger network, highlighting the metabolite specificity of these rQTL (**Figure 4**f).

Interpreting the biology underlying the rQTL between amino acids from these two loci is relatively straightforward: Asparagine and threonine are separated by 6 reactions from each other according to KEGG (**Supplementary Table 10**), while glutamine and asparagine are involved in a single reaction (EC number: 6.3.5.4) and asparagine and threonine are 2 reactions away from each other (**Supplementary Table 10**). Therefore, it is likely that the altered enzymatic function through the missense variant changes substrate availability, explaining the metabolites in the ratio. However, according to the SwissProt database, neither ASPG (”Q86U10”) nor GLS2 (”Q9UI32”), have catalytic effects on n-acetylated metabolites, making the interpretation more difficult [34]. Here, we propose three hypotheses: i) Indirect regulation between the amino acid and its n-acetylated form. ii) unexpected catalytic function of the enzymes potentially through the missense variant or iii) unexpected measurement interactions between the metabolites across these clusters.

### Using rQTL to instrument proteins abundances for drug target analysis

The rQTLs found in this study can be used to proxy the activity of the enzyme that underlies the metabolic reaction [35]. For example, we identify a missense variant (rs77863699) in Acyl-CoA Synthetase Medium Chain Family Member 2B (*ACSM2B*) in LD *R*^2^ = 0.95 with the novel rQTL between salicylic acid and salicylurate (rs9922704, rQTL *P* = 4 *·* 10*^−^*^35^, *P_gain_* = 10^29^) (**Supplementary Table 5, 6**). Salicylic acid is a xenobiotic and a metabolite of aspirin. The breakdown of salicylic acid happens through glycine conjugation into salicylurate [36]. Glycine conjugation can be used to detoxify xenobiotics, allowing their excretion through the kidneys, which is a two-step reaction, by first bonding salicylic acid to acetyl-CoA using the *ACSM2B* enzyme (Reactome reaction ID: R-HSA-159567), and then conjugating glycine [37]. These metabolites did not pass significance thresholds in the source study for marginal associations (minimum *P* = 6 *·* 10*^−^*^6^), and indicate that this ratio can proxy the activity of the *ACSM2B* enzyme, which is not measured in the Interval, UKB-PPP nor deCODE proteomics studies [4, 38, 39].

It is possible to generalize our rQTL findings to (pseudo) enzymatic activity. For this, we combine the 104 genes containing a missense variant in strong LD *R*^2^ *>* 0.8 with an rQTL, and the 29 genes identified through eQTL MR, resulting in a total of 125 unique genes (**Supplementary Table 6, 7**). We are aware that these rQTLs do not strictly instrument protein abundance, however the enzymatic enrichment (**Figure 2c**), and the limited false positive rates of MR-link-2 [27, 40] suggest that these rQTLs likely serve as a surrogate for the genes’ enzymatic function. Interestingly, when we compare these genes to previously measured pQTL proteins from the Interval, UKB-PPP and deCODE proteomics QTL studies [4, 38, 39], we are able to proxy 91 (72%) otherwise unmeasured proteins, including 5 Cytochrome p450 genes: *CYP2C19*, *CYP2C8*, *CYP2C9*, *CYP4B1* and *CYP4F2* (**Supplementary Table 6,7**). Four genes that are uniquely ’instrumentable’ through our rQTLs also contain approved drug targets according to OpenTargets [41], which opens new avenues to use rQTLs to instrument otherwise unmeasured protein levels/enzymatic activities, showing that rQTL analysis can be informative for drug target identification and validation [35].

### Using rQTL to identify driver genes outside whole blood

Metabolism does not always occur in the blood, and therefore, it is likely that the underlying genes cannot be instrumented through whole blood eQTL or pQTL [4, 26, 42, 43]. We, therefore, tried to link our blood metabolite-based rQTLs with tissue specific gene expression across 48 tissues from the GTEx consortium [44] (**Methods**). To this end, we performed 554,898 *cis*-eQTL to rQTL to MR tests, using an instrument selection threshold of 5*·*10*^−^*^8^, finding 727 rQTL that can be explained by tissue-specific gene expression (MR-link-2 *P <* 9 *·* 10*^−^*^8^), comprising 250 gene-tissue combinations, 41 out of 48 tissues and involving 46 unique genes (**Supplementary Table 12**). We combine these tissue-specific causal gene findings from whole blood eQTL genes and genes identified from missense variants. This allows us to identify 26 rQTLs that are eQTLs exclusively in non-blood tissue. Thus, inclusion of the GTEx data allows for the assignment of a causal gene for another 121 individual rQTLs across 22 LD based clusters (**Supplementary table 12**).

Through this multi-tissue causal inference, we identify a causal estimate between the expression of the Electron Transfer Flavoprotein Dehydrogenase (*ETFDH*) gene and 12 metabolite ratios. Only through this skeletal muscle specific expression of *ETFDH* is it possible to identify Bonferroni significant MR-link-2 estimate onto 12 ratios of metabolites that fully comprise carnitine containing metabolites (**Supplementary Table 5**). Carnitines covalently bind fatty acids for transport to the mitochondria, where fatty acids are used for beta oxidation and produce ATP through oxidative phosphorylation in which *ETFDH* is an essential protein [32]. This eQTL for the *ETFDH* muscle expression was not linked to any other molecular trait (via blood pQTLs, non-muscle eQTLs). *ETFDH* is under selective pressure, as recessive mutations cause glutaric acidemia II (MIM number 231680) [45, 46]. The relationship between *ETFH* and carnitines can be further explained through the glutaric acidemia II disorder, which is treated by supplementation of L-carnitines [45].

These results highlight that rQTLs allow for the identification of cross-tissue biology, which can hint to which tissue may be relevant for the rQTL mechanism, but perhaps more importantly, can be used to instrument genes that are otherwise only functional in difficult-to-access tissues, which can be helpful in drug target analysis [35].

### rQTL to identify causal genes for disease and their source tissue

Up until this point, our analysis has been directed at the identification of the underlying causes of the rQTLs. Illustrated by the example of glutaric acidemia II, rQTLs can also be used to instrument disease biomarkers and reveal new mechanisms, potentially informing treatment opportunities and drug development. To this end, we analyzed the causality between the metabolite ratios found in this study and 327 disease relevant traits compiled by Kurki *et al.* [47].

Using a Bonferroni threshold of 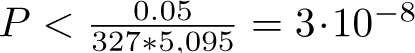, we identify 961 Bonferroni significant MR estimates across 25 complex traits and 625 metabolites ratios. The most common trait is pure hypercholesterolemia (ICD10: E78.00), which is consistent with the abundance of rQTLs regulating ratios involving lipids (**Supplementary Table 13**) [3].

Remarkably, we identified 26 metabolite ratios where their rQTLs had a significant MR estimate onto malignant bladder neoplasms. These MR hits were distributed across 5 LD independent rQTL clusters in 4 loci (**Supplementary Table 5, 13**). Upon inspection of the 15 metabolites that comprise these ratios, we could map 7 metabolites to the Wikipathways caffeine pathway (”WP3633”) [14] (**Figure 5a**). Moreover, 3 out of 4 loci mapped to 3 out of 4 enzymes in this pathway: CYP1A2, CYP2A6 and NAT2, missing only the XDH enzyme (**Figure 5a,b**). The remaining locus that did not contain enzymes in the caffeine pathway showed a single novel rQTL for the ratio between metabolites that are in the caffeine pathway: 7-methylxanthine and 3,7-dimethylurate (rs115430557, *P* = 3.0 *·* 10*^−^*^14^), and is located within the glutathione S-transferase (GST) cluster of genes at 1p13.3 (**Figure 5b**). GST enzymes are involved in the detoxification of electrophilic compounds. We performed MR to understand which genes are important across these loci, where we only identified significant MR estimates for the *GSTM* locus. Causal estimates based on eQTLs pinpointed *GSTM1*, *GSTM2*, and pQTL-based MR revealed GSTM4, which had the strongest association (*P* = 2.6 *·* 10*^−^*^17^) (**Figure 5c**) (**Supplementary Tables 7,8**).

**Fig. 5.**
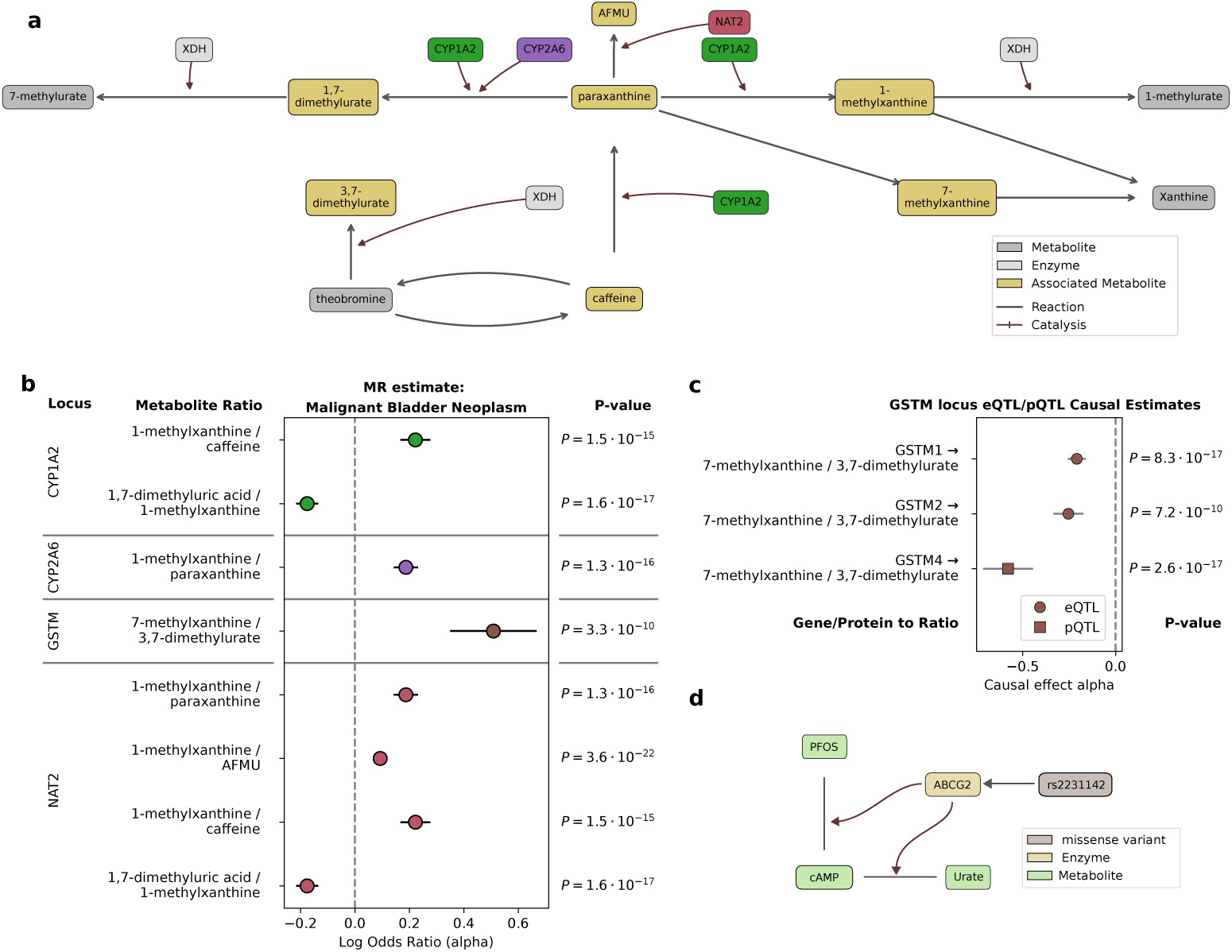
a) The WikiPathways human caffeine pathway (”WP3633”) with metabolites and catalysts. Nodes are colored depending on if they were a constituent metabolite of an rQTL that passes MR significance for causal to malignant bladder neoplasms. b) Causal effect estimates between metabolite ratios of metabolites that are fully present in the caffeine pathway (exposure) onto malignant bladder neoplasm, passing Bonferroni significance. Error bars indicate the 95% confidence interval of the MR-link-2 estimate. c) *GSTM1*,*GSTM2*, and GSTM4 are the only molecules that have significant MR causal estimates that link the metabolite ratios estimated to be causal for bladder cancer. The GSTM locus contains detoxifying enzymes that are not found in the caffeine pathway, but are related to the caffeine metabolite through as the metabolites in the ratios are present in the pathway. These exposures are causal to the rQTL between 7-methylxanthine and 3,7-dimethylurate. Error bars indicate the 95% confidence interval of the MR-link-2 estimate. d) The emerging pathway instrumented by variants in the *ABCG2* locus: The *ABCG2* loss of function missense variant, rs2231142, is an rQTL for the adenosine cyclic monophosphate (cAMP) to perfluorooctanesulfonate (PFOS) and cAMP-to-urate ratios (lines between metabolites).

The mapping of these rQTL to caffeine enzymes and malignant bladder neoplasms do not necessarily point to a causal relationship between caffeine metabolites and cancer, but rather capture the detoxifying effects of the NAT2, CYP1A2, CYP2A6, GSTM1, GSTM2 and GSTM4 enzymes [48–51]. Indeed, all the loci we identified through the significant link with bladder cancer contained enzymes (CYP1A2, CYP2A6, NAT2 and GSTM1) have been described affect the breakdown of smoking related metabolites [52–55]. The hypothesis that these rQTL proxy detoxifying effects is further strengthened by the observation that there is exclusive complex trait that is co-significant with malignant bladder neoplasms, namely malignant lung neoplasms from 2 metabolite ratios to complex trait MR associations (**Supplementary Table 13**). Similar to malignant bladder neoplasms, the largest risk factors for developing malignant lung neoplasms are due to exposures like smoking [56]. These results reiterate that rQTLs can instrument disease relevant enzymes which are otherwise difficult to detect using protein measurement based studies, as 4 out of the 6 enzymes reported in this analysis were not found in any of the large pQTL studies we queried [4, 38, 39]. We found a further link between six rQTLs, located in the Stearoyl-CoA Desaturase (*SCD*) locus, linked to ratios between lipids that were MR-significant for ischaemic heart disease, a further two of these metabolite ratios were also MR-significant for coronary atherosclerosis (**Supplementary Table 13**). *SCD* encodes for an enzyme that catalyzes the desaturation of fatty acids, specifically palmitoyl- and stearoyl-CoA, introducing a single saturation into an unsaturated fatty acid [57]. High SCD activity is considered favorable for cardiovascular health, and its activity can be clinically proxied through a metabolite ratio between palmitoate (C16:0) and palmitoleic acid (C16:1n7) [57, 58]. Reassuringly, we found an rQTL for the same ratio palmitoate (C16:0) and palmitoleic acid (C16:1n7), providing a genetic association that proxies this activity. Furthermore, all ratios associated with this rQTL include a pair of an unsaturated fatty acid, and a saturated fatty acid (**Supplementary Table 5, 11**), suggesting that these rQTLs actually represent SCD activity.

Unfortunately, despite these rQTLs mapping to the *SCD* locus, we do not find MR evidence using pQTLs, eQTLs or tissue specific eQTLs as exposures. However, we do identify *SCD* effects in adipose tissue at the more lenient Benjamini-Yekutieli false discovery rate of 0.05 (*P* threshold = 5.5 *·* 10*^−^*^5^) [59]: The most significant MR estimate is between *SCD* gene expression in the Adipose Visceral Omentum tissue (*P* = 4.4 *·* 10*^−^*^7^), as well as in Adipose Subcutaneous tissue *P* = 2.0 *·* 10*^−^*^5^) on the palmitate-to-palmitoleic acid ratio (**Methods**) (**Supplementary Table 12**).

### Proposing *ABCG2* as a PFOS transporter

Perhaps the most notable rQTL of this study is rs149027545 (*P* = 6.3 *·* 10*^−^*^19^) associated with the adenosine cyclic monophosphate (cAMP) to perfluorooctanesulfonate (PFOS) ratio. Notably, this rQTL is in full LD (*R*^2^ = 1.0) with a missense variant (rs2231142) in the ATP Binding Cassette Subfamily G Member 2 (*ABCG2*) gene. Individually, the metabolites in this ratio do not pass the Bonferroni significance threshold (*P <* 4.5 *·* 10*^−^*^11^) of the source study (min mQTL *P* = 1.0 *·* 10*^−^*^10^), making it novel [3] (**Supplementary Table 5**).

PFOS is one of the man-made ’forever chemicals’, which accumulate in the environment and are difficult to degrade [60]. High concentrations of PFOS and other per-and polyfluoroalkyl (PFOA) substances have adverse effects on human health, leading to thyroid disease as well as kidney- and testicular cancer [61]. PFOA are excreted in humans through menstrual blood and the intestines [61, 62]. PFOA excretion half-lives have been estimated to be in the order of magnitude of years [63]. To our knowledge, the mechanism of PFOS excretion has not been fully elucidated.

Our results suggest that PFOS is excreted through *ABCG2*, likely into the intestines in a cAMP dependent way. *ABCG2* is a urate transporter, which is expressed in the intestines, and considered a causal risk factor to gout [64]. We provide further support to our hypothesis: First, we additionally found an rQTL for the cAMP-to-urate ratio at rs45499402 (*P* = 6.3 *·* 10*^−^*^19^, LD *R*^2^ = 0.993 with the missense variant rs2231142) (**Figure 5d**). Furthermore, we identify a causal relationship between the cAMP-to-urate ratio and Idiopathic Gout (MR-link-2 *P* = 2.9 *·* 10*^−^*^11^) using rQTL instruments from the *ABCG2* locus. The rQTL missense variant in *ABCG2* results in a glutamine to lysine substitution that reduces the urate excretion rate by 54%, making it a loss of function variant [64, 65]. Additionally, the relationship between *ABCG2* and PFOS can be further supported by epidemiological observation that PFOS and urate levels are positively correlated [66].

In the context of *ABCG2* it is also possible to explain the other trait in the ratio: cAMP is a signaling molecule that responds to changing conditions in the cell [67, 68]. Both the relationship between PFOS to cAMP and urate to cAMP can be explained through *ABCG2*, which excretes urate, but also other xenobiotics in a cAMP dependent manner [69, 70]. *ABCG2* contains a cAMP-sensitive promoter, indicating a dose-response relationship between *ABCG2* abundances and its transport activity [67, 68] (**Figure 5d**).

## Discussion

In this work, we propose a methodology to estimate ratio quantitative trait loci (rQTLs), using only publicly available summary statistics from an mQTL study, bypassing the need for individual level data. This approach allows for the identification of rQTLs between hundreds of thousands of ratios within modest computational requirements. We validated the method extensively on rQTL summary statistics derived from individual level data and find generally good agreement in terms of effect size and significance. We consider our methodology conservative, as the large majority of rQTLs identified in the validation set were also confirmed in the source study.

Next, we applied the methodology to all pairwise ratios of 1,091 metabolites (594,595 ratios), and found 5,095 ratios that pass stringent Bonferroni corrected *P* value and *P_gain_* thresholds [15]. These rQTLs are enriched for enzymes when selecting genes using (in order of increasing enrichment) closest genes, missense variation, eQTL Mendelian randomization (MR) and pQTL Mendelian randomization (MR). Furthermore, metabolite ratios with rQTLs have shorter reaction distances than metabolite ratios that do not have rQTLs. Approximately 21% of the loci identified through our methodology are novel. I.e. these were not identified in the marginal association analysis at Bonferroni significance threshold in the source study [3], highlighting that testing meaningful ratios can lead to new discoveries. For instance, the rQTL estimation can also be applied to gene expression, protein abundance or methylation phenotypes.

We find that the rQTLs found in our study often capture enzymatic activity: We recapitulate fatty acid desaturation and elongation through the *FADS2* and *ELOVL2* loci, as well as substrate specific rQTLs between glutaminase and asparaginase, where amino acids have exclusive rQTLs with other amino acids. The same holds for the n-acetylated metabolites, which have rQTLs only when we test ratios with one another. The instrumentation of enzyme activity can be relevant for researchers that investigate drug-targets: We are able to instrument the *ACSM2B* enzyme which is involved in the detoxification of salicylic acid, a metabolite of aspirin. Across the genes identified by this rQTL study, 72% are not measured in pQTL studies, showing the potential for rQTL to instrument difficult-to-measure proteins.

We interpret a gene found through an rQTL and pQTL differently. Namely that rQTLs may represent the activity of an enzyme, while pQTLs may rather reflect steady-state abundance of the proteins, and not necessarily catalytic activity. Moreover, rQTLs are more likely to reflect activity in a blood-independent tissue. This hypothesis is supported by several examples when rQTLs colocalise with (muscle or adipose) tissue-specific expression levels obtained from GTEx eQTLs [44].

rQTLs can be even more directly informative for human health: We identified causal relationships between metabolite ratios in the caffeine pathways and malignant bladder neoplasms as well as lung neoplasms. These causal relationships are not necessarily due to caffeine metabolites, but rather serve as a surrogate for the relevant xenobiotic detoxifying enzymes. Similarly, we saw an rQTL likely proxying adipose tissue specific *SCD* expression that is relevant for cardiovascular health.

Finally, we identified a novel rQTL near the *ABCG2* gene that turned out to be relevant for the excretion of PFOS. This result was unexpected and may provide an explanation for how humans excrete these difficult to degrade ’forever chemicals’. We find this result plausible due to the clear cut loss-of-function effect of the missense variant that is tagged by the rQTL [64, 67], combined with the cAMP-active element in the gene promoter [67], together with the epidemiological findings that urate and PFOS concentrations are correlated [66]. It is, however, necessary to perform functional follow-up experiments before any proof can be given that ABCG2 is the transporter through which PFOS is excreted.

Not only can blood rQTLs be used as a proxy for protein/enzyme activity, but sometimes can even proxy activities in other tissues. Despite the richness of biobank scale measurements, molecular measurements are limited to minimally invasive tissues for biopsy such as blood plasma, which can be problematic when targeting a disease that is not blood-relevant [42, 71]. Metabolites are among the smallest molecules that can be reliably measured, and, it stands to reason that small metabolites are able to cross tissue boundaries to the plasma more easily. This is the case for the blood-brain barrier, which has been shown to be more permeable for smaller molecules[72].

The methodology outlined in this study to identify rQTL is not limited to metabolites or strictly molecular traits. Our methodology requires the phenotypes of interest to have been transformed via a function that is similar to the logarithm. Many concentration-based traits (metabolites, proteins, transcripts) are log transformed, hence these are the primary targets to be tackled in the future. The potential to increase the number of findings by approximately 20 % by considering ratios is a compelling prospect.

One interesting aspect underlying many rQTLs is that one locus can be linked to multiple ratios. This can increase robustness of the associations, as it allows us to leverage multiple proxies for the same protein abundance/activity. This can be especially valuable when considering metabolite ratios as potential biomarkers. Using multiple ratios that are associated to the same locus is likely to give the potential biomarker more weight in follow-up.

This study has several limitations: even though we reidentify many catalyzing enzymes, there is always a possibility that rQTLs estimated in this way contain false positives [20]. We tried to reduce this as much as possible by (i) only reporting rQTL at a stringent Bonferroni threshold; (ii) requiring strong *P_gain_*; (iii) only reporting missense genes in strong linkage disequilibrium (LD); and (iv) using an MR method with strong type I error control. Nonetheless, across all comparisons, we expect to find some false positives.

Even though rQTL are able to identify enzymatic function, it is still unclear to what extent. There are still a lot of unknowns as to what a metabolite ratio can represent, but using the methodology presented in this study, and combined with the increasing sample sizes of molecular QTL studies, we will increase our understanding of rQTL relevance to human biology.

## Methods

### Summary statistics of metabolite QTL studies

The summary statistics that we used to create ratio summary statistics are from the European component of the Chen *et al.* study [3] (GWAS Catalog Identifiers: GCST90199621-90201020). The 1,091 metabolites in Chen *et al.* were harmonized to Human metabolite database (HMDB) identifiers by taking the Chen et al. harmonization tables, as well as through the procedure previously described by van der Graaf *et al.* [27].

We used classically calculated ratio summary statistics from Chen et al. (2023) [3] as the source of ground truth in validating the derived ratio summary statistics methodology. These were also downloaded from the GWAS catalog using GWAS Catalog Identifiers GCST90199621-90201020. Of the 309 available ratio summary statistics, 299 corresponding to ratios of only two metabolites were selected. The 10 ratio traits with more than two underlying metabolites were excluded.

### Summary statistics harmonization

In this work, we harmonized summary statistics in the following way: First, if necessary, we lifted over summary statistics into human chromosome build 37 using UCSC’s liftover tool (https://genome.ucsc.edu/cgi-bin/hgLiftOver) combined with their chain files (https://hgdownload.soe.ucsc.edu/downloads.html). Then, we include SNP variants that have LD information available, by overlapping the variants (based on chromosome, position and alleles) present in the summary statistics file with the variants in our LD reference (UK10K) [73]. To improve robustness and to minimize outliers, we selected variants with a minor allele frequency (MAF) higher than 1%.

### Derivation of ratio summary statistics

Based on the derivations of Karsten Suhre [19], it is possible to derive summary statistics of a ratio phenotype [19]. The summary statistics of a ratio phenotype is listed below:

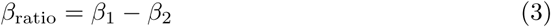

Where *β*_ratio_ is the genetic effect size of the ratio phenotype, *β*_1_ is the effect size of the nominator phenotype and *β*_2_ is the effect size of the denominator phenotype. As both *β*_1_ and *β*_2_ have a standard error associated to them (*σ*_1_*, σ*_2_ respectively), it is possible to calculate the standard error of *β*_ratio_ using the following formula.

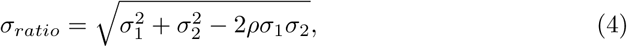

Where *ρ* is a correlation term of the two phenotypes. In the development of this methodology, we tested two different ways in which this correlation term can be determined:

1. The cross-trait LD score regression intercept (estimating phenotypic correlation while accounting for sample overlap)
2. The correlation value for which the median test statistic follows the null distribution

The selection of the *ρ*_1,2_ values was done in the following way: i) The LD score regression intercept was used as *ρ*_1,2_, which came from the LD score regression software (https://github.com/bulik/ldsc) [24]. ii) We determine the *ρ*_1,2_ by searching for the optimal correlation for which the median *χ*^2^ value of a summary statistics is closest to its expected 0.455, also known as the genomic inflation factor [74].

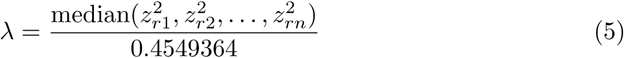

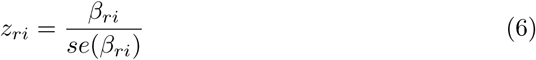

We optimize for the *ρ*_1,2_ factor using a brentq root-finding algorithm implemented in scipy, which identifies the *ρ*_1,2_ factor that identifies the best fit to ensure there is no genomic inflation or deflation. Code for this procedure can be found at https://github.com/adriaan-vd-graaf/sumstats-ratio-qtl

### Identification of Independent Genetic Loci via Clumping

We performed clumping using plink (v1.90b7.3) [75], using the UK10K cohort as an LD reference [73]. We applied this procedure for two purposes:

1. To identify independent SNPs inside an rQTL locus, we identified all SNPs that passed our significance thresholds: *P <* 8.4 *·* 10*^−^*^14^ and *p_gain_ >* 5, 945, 950. To ensure that the SNPs were highly independent, we selected clumped top variants at a stringent LD threshold of *R*^2^ *>* 0.0001.
2. To select independent instruments for MR, associated regions are identified through clumping at a *P <* 5 *·* 10*^−^*^5^ for blood molecular traits onto metabolite ratios and *P <* 5 *·* 10*^−^*^8^ for GTEx eQTL onto metabolite ratios and comparisons between metabolite ratios and complex traits. Clumping was performed used the default parameters of MR-link-2 with a clumping window of 250 kb and at a less stringent LD threshold of *r*^2^ *<* 0.01 [40].

The *P_gain_* statistics is used to ensure that the ratio QTL is substantially more significant than the underlying metabolites. We calculated *P_gain_* using the equation in Peterson *et al.* [15]: 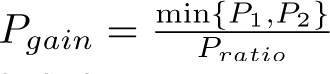 where *P*_1_ and *P*_2_ are the P values of two SNP-metabolite associations, and *P_ratio_* is the *P* value of the ratio association. When ratios between two random metabolites are tested, a *P_gain_* of 10 is expected to be observed in 5% of the cases, roughly representing a P value threshold of 0.05 [15]. To ensure a Bonferroni significant *P_gain_*, we multiply our threshold with the number of tests performed to identify rQTL making our threshold 5,945,950.

### Validations of the loci found by the original study, and the derived ratio methodology

To validate our methodology for derivation of the ratio summary statistics using the genomic inflation factor method, we clumped all metabolite pair ratios that Chen *et al.* found at their mQTL Bonferroni significant threshold (*P <* 4.6 *·* 10*^−^*^11^). We determine how often we re-identify an rQTL of the loci between the rQTL published by Chen *et al.*, and the ratios calculated in this study. Some rQTLs were identified by our methodology and not by Chen *et al.*, these do not pass significance in either the mQTLs or the rQTLs (**Supplementary Table 4**).

### Derivation of newly-found ratio summary statistics

We generate summary statistics for metabolite ratios, using the genomic inflation factor control method to estimate *ρ*, based on good results in the validation set (**Supplementary Table 1**). From 1,091 metabolite summary statistics, we calculate (1, 091 *·* 1, 090)*/*2 = 594, 595 ratio summary statistics. To identify potentially interesting ratio-associated SNPs, we selected ratios that had at least one SNP with a Bonferroni-corrected *P* value below 8.42 *·* 10*^−^*^14^ = 5 *·* 10*^−^*^8^*/*594, 595 and a Bonferroni-corrected *P_gain_* greater than 10 *·* 594, 595. SNPs meeting both the Bonferroni-corrected *P* value and *P_gain_* thresholds were then clumped. For each selected ratio, we defined *cis*-regions as 250 kilobases (kb) around the lead clumped SNP. As an additional filter, we only select the regions in which the top rQTL has a significant *P_gain_* considering all the SNPs in that 250kb region, using the summary statistics within these regions for subsequent analyses. We refer to these Bonferroni-significant *P* value and *P_gain_* lead SNPs as rQTLs.

### Definition of novelty based on locus

An rQTL was designated ’novel’ if none of the constituent individual metabolites passed the Bonferroni significance threshold in the same locus in the source study. We compare the lead rQTL variant with a 250 kb window and call the rQTL novel when there is no constituent metabolite with an individual *P* value smaller than 4.6 *·* 10*^−^*^11^ = 5 *·* 10*^−^*^8^*/*1091.

### Clustering of rQTLs

Clumped rQTLs were clustered based on their linkage disequilibrium (LD). For this, we constructed an undirected graph where each rQTL SNP is a node. An edge is drawn between each pair of nodes if and only if the pairwise LD, measured as *r*^2^, was greater than 0.1 (**Supplementary Table 5**). Finally, we clustered this graph using the infomap python API https://mapequation.github.io/infomap/python/ [76].

### Closest gene and missense variants in LD with rQTLs

To match the rQTL to genes, we performed closest gene mapping and identified missense variants in LD with the rQTL. The closest genes were identified using Ensembl gene mapping [77]. We identified variants in strong LD *≥* 0.8 with our rQTLs in the following way: The rQTLs were provided as the index SNPs for this analysis. The resulting list of LD proxies was then annotated for potential functional consequences using the Ensembl Variant Effect Predictor (VEP) command line package (version [113.0]) [25]. We only kept variants predicted to be missense mutations (**Supplementary Table 6**).

### Mendelian randomization

To further identify genes for gene expression and protein levels, we used MR-link-2 as the main Mendelian randomization (MR) method in this study [27]. This *cis*-MR method is appropriate because of the *cis-* (single region) nature of the rQTL under study and the limited Type 1 error of the MR-link-2 method. Additionally, the MR-link-2 package also performs other *cis* MR analysis as well as coloc analysis. The implementation for MR-link-2 can be found at (https://github.com/adriaan-vd-graaf/mrlink2). We used MR-link-2 with standard settings, using the UK10K panel as an LD reference [73].

### Summary statistics whole blood of eQTLs

To estimate causality from gene expression onto the metabolite ratios, we performed MR on mRNA transcript expression QTL (eQTL) data from the eQTLGen Consortium [26]. For this study, we used *cis*-expression quantitative trait loci (eQTL) summary statistics from eQTLGen phase 1, which includes association results for 19,960 genes across 31,684 blood samples. These summary statistics were obtained from the eQTLGen website (https://www.eqtlgen.org).

### Summary statistics of whole blood protein QTLs

To estimate causality from protein abundance onto the metabolite ratios, we performed MR on protein QTL (pQTL) data from the UK Biobank Pharma Proteomics Project (PPP) [4](Downloaded from: https://doi.org/10.7303/syn51364943). The PPP study contains genetic associations for 2,923 plasma proteins across 54,219 individuals. The summary statistics and metadata were downloaded from the respective resource and harmonized using the UniProt identifiers provided by the study. For subsequent *cis*-QTL analysis, we focused on SNPs located within a *±*1 megabase (MB) window of each protein-coding gene’s transcription start site (TSS) based on the Ensembl gene mapping [77].

### Summary statistics of multi tissue eQTL

To interrogate the role of tissue-specific gene expression, we estimated causality from tissue specific gene expression onto metabolite ratios. We performed MR using the the summary statistics from the Genotype-Tissue Expression (GTEx) project [44]. We utilized *cis*-eQTL data from GTEx release v8, and we included association results for gene expression across 48 different human tissues. This dataset was incorporated to determine the impact of gene expression in a tissue-relevant context, complementing the blood-specific analysis from eQTLGen. The summary statistics were downloaded from the European Bioinformatics Institute (EBI) public FTP server (https://ftp.ebi. ac.uk/pub/databases/spot/eQTL/imported/GTEx V8/ge/).

### Summary statistics of FinnGen complex traits

For our analyses, we used publicly available genome-wide association study (GWAS) summary statistics from the FinnGen Data Freeze 12 meta-analysis. This dataset incorporates data from the Finngen cohort (R12), the UK Biobank (UKBB), and the Million Veteran Program (MVP). The MVP data included participants from Admixed American, African, and European ancestries. The phenotype harmonization and meta-analysis procedure are extensively described according to the study documentation. [47]. We downloaded the meta-analysed summary statistics following the instructions on https://www.finngen.fi/en/access results.

### Estimating causality between molecules and metabolite ratios and between metabolite ratios and disease

To identify causality between whole blood protein and expression levels, using pQTLs and eQTLs as instruments and metabolite ratios as outcome, we clumped eQTL and pQTL variants using a significance threshold of *P <* 5 *·* 10*^−^*^5^ to identify associated regions. We subsequently tested for causality using MR, and identified significant causal estimates at a Bonferroni multiple testing threshold, where the number of tests is used as a correction factor (**Table 1**).

**Table 1.**
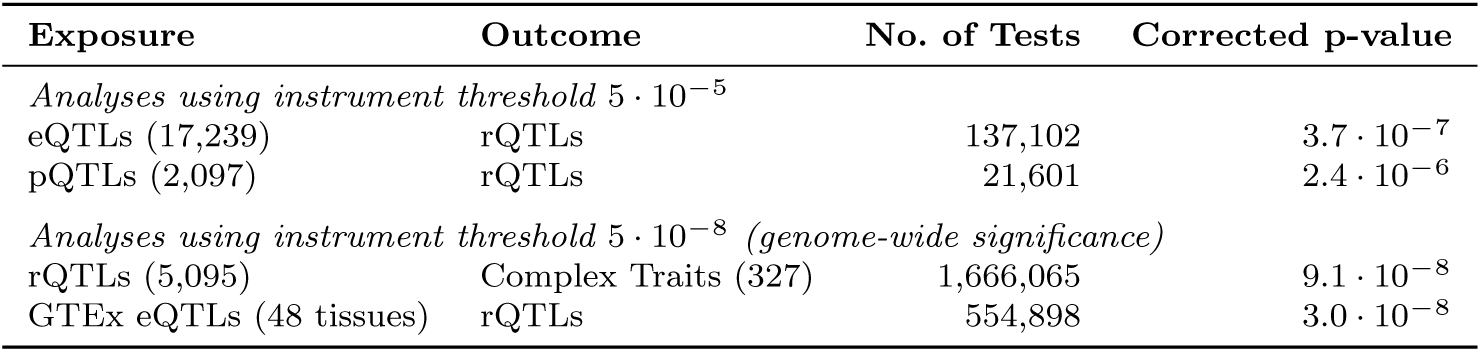
Mendelian randomization tests and multiple testing thresholds

| Exposure | Outcome | No. of Tests | Corrected p-value |
| --- | --- | --- | --- |
| *Analyses using instrument threshold $5 \cdot 10^{-5}$* |  |  |  |
| eQTLs (17,239) | rQTLs | 137,102 | $3.7 \cdot 10^{-7}$ |
| pQTLs (2,097) | rQTLs | 21,601 | $2.4 \cdot 10^{-6}$ |
| *Analyses using instrument threshold $5 \cdot 10^{-8}$ (genome-wide significance)* |  |  |  |
| rQTLs (5,095) | Complex Traits (327) | 1,666,065 | $9.1 \cdot 10^{-8}$ |
| GTEx eQTLs (48 tissues) | rQTLs | 554,898 | $3.0 \cdot 10^{-8}$ |

To identify tissue-specific gene expression that is estimated to be causal to a metabolite ratio, using the GTEx eQTLs, we performed the same MR analysis, but at a more stringent *P <* 5 *·* 10*^−^*^8^ instrument selection threshold. For analyses of *cis*-pQTL and *cis*-eQTL (all tissues including whole blood) effects on rQTLs, comparisons were restricted to instances where the respective *cis*-regions overlapped with the rQTL regions that were found.

To identify the downstream effects of the rQTL significant metabolite ratios that we identified in this study, we perform MR using the metabolite ratios as exposures, and the complex traits reported by the FinnGen MVP and UKBB meta-analysis as the outcome. For this, we perform clumping in the rQTL using a *P* value threshold of *P <* 5 *·* 10*^−^*^8^

### Enzymatic enrichment of rQTLs

To investigate whether rQTLs are enriched in genes encoding enzymes, we first annotated the mapped genes for enzymatic activity using the UniProt database [28]. Selecting for reviewed proteins that are specific to humans and have catalytic activity https://www.uniprot.org/uniprotkb?query=%28reviewed%3Atrue%29+AND+%28organismid%3A9606%29&facets=proteinswith%3A13 (accessed September 27, 2025) [28]. We enriched 4 gene mapping methods for enzymes: closest genes, missense genes, eQTL MR and pQTL MR. To enrich the closest genes to the clumped SNPs and genes found through missense variant mapping, we contrasted these gene sets against all human protein coding genes (Ensembl biomart, downloaded 26th of July 2025). To enrich the genes found by the eQTL MR and pQTL MR methods, we contrasted the genes found through MR with the genes that have an MR estimate, but did not reach significance.

### Creation of the network graph with minimum distances

To compare metabolite ratios based on the reaction distance between their underlying metabolites, we calculated the minimum reaction distance using information from our KEGG, metacyc and wikipathways network graphs [27]. We then categorized ratios into two groups: those with variants meeting significance thresholds for *P* -values (*<* 5 *·* 10*^−^*^8^*/*594, 595) and *P_gain_* (*>* 10 *·* 594, 595), and those without rQTLs. Using a lookup in our minimum reaction distance graph, we identified reaction distances for 12,438 ratios without rQTLs and 117 ratios with rQTLs. (**Supplementary Table 10**)

### Mapping saturations and number of carbons of fatty acids

We mapped saturations of fatty acids by counting the number of carbons and double bonds in the chain as defined by the fatty acid identifier using regular expressions that can be found at github.com/adriaan-vd-graaf/sumstats-ratio-qtl (**Supplementary Table 11**). We did not include fatty acids that had multiple carbon chains, or where there was ambiguity in the number or placement of double bonds.

We performed enrichment analyses for the absence of zero double bonds at the *FADS* locus. Here, we compared the number of ratios with zero double bond difference to the number of ratios with a non-zero double bond difference. We counted these occurrences for fatty acid ratios inside the *FADS* locus, and for fatty acid ratios outside the fads locus

Additionally, we performed enrichment analysis for the presence or absence of an even or odd number of carbons differences between fatty acids in the *ELOVL2* locus. Here, we counted the number of even and uneven carbon chain differences between monoacylic fatty acids, and compared these numbers between ratios inside the *ELOVL2* locus and ratios outside the *ELOVL2* locus

#### Author contributions

A.vd.G. and Z.K. conceptualized the study, A.vd.G. and S.R. performed data curation and analysis. S.R., A.vd.G. and Z.K performed formal analysis, including statistical, and computational techniques to synthesize and analyze data. Z.K. acquired funding. A.vd.G., S.R., and Z.K. developed and designed methodology, Z.K. provided = resources. A.vd.G. and S.R. performed software development. A.vd.G. created visualizations and wrote the original manuscript draft. A.vd.G., N.G., S.R. and Z.K. reviewed, and edited the manuscript.

#### Data and Code availability

The code that was used to calculate the metabolite ratios is published at github. com/adriaan-vd-graaf/sumstats-ratio-qtl. The summary statistics of the rQTL regions that are built in this study can be downloaded from metabolite-ratio-app.athirtyone. com/. The genotype information underlying the LD matrices for the UK10K data resource were downloaded from the EGA under accession IDs EGAD00001000740 and EGAD00001000741. As this is individual level genotype data, a data access agreement is required for access. Conditions for this data access agreement can be found at https://www.uk10k.org/data access.html.

## Data Availability

All data produced in this work are contained in the manuscript, the supplementary information or are available online at metabolite-ratio-app.unil.ch

https:///metabolite-ratio-app.unil.ch

## Acknowledgements

We thank all study participants for their altruistic donations of their data. We would further like to thank all collaborators (eQTLGen, UKB-PPP, GTEx, FinnGen, UK Biobank, and the Million Veterans Cohort study) that have provided data resources for use in this study. We would also like to thank in particular all the authors of the Chen *et al.* study, without whom this study would have looked very different. Z.K. is funded by the Swiss National Science Foundation (SNSF 315230-219587).

## Supplementary Table legends

**Supplementary Table 1**

The Metabolites and metabolite ratios (”type”) used in this study (”Accession”, ”reportedTrait”), the ancestry and sample size of (”discoverySampleAncestry”) and the download location (”summaryStatistics”).

**Supplementary Table 2**

Comparison between genomic inflation factor and LD score intercept term. For all the ratios tested in this study (”numerator accession”, ”denominator accession”), the estimates of the LD score regression intercept value (”cor-relation term estimated by ldsc intercept”), combined with the estimate of the correlation term where the genomic inflation factor is controlled. (”correla-tion term estimated by genomic inflation factor control”)

**Supplementary Table 3**

Validations that were performed between classically derived rQTL, and the rQTL identified in the method used here. We compared effect sizes (betas) and Z scores (”Comparison type”) for 299 classically derived ratios (”Accession”), at two different P value filter thresholds 1.0 (unfiltered) and marginal significance P ¡ 0.05. For each of these categories, we performed linear regression (slope and intercept), Pearson correlation and Spearman correlation.

**Supplementary Table 4**

Validation of clumped variants (at *P <* 4.6 *·* 10*^−^*^11^) found in the validation set from the Chen *et al.* study, the underlying mQTL, and the rQTL found through our methodology.

**Supplementary Table 5**

All the rQTL identified using our methodology across all the metabolite ratios that were tested. We include information for the ratio (including the reaction distance), the association of the significant clumped genetic variant, the information of the constituent metabolites as well as a summary of the gene mapping methods that passed our significance thresholds.

**Supplementary Table 6**

Missense variants found located close to the clumped rQTL variants. For each ratio (”ratio accession”, ”num name”, ”den name”) with an rQTL (”rsid”, ”infomap cluster”) we mapped missense variants to genes (”missense gene”, ”mis-sense rsid”) when their LD *R*^2^ was larger than 0.8 (”ld with clumped variant”)

**Supplementary Table 7**

Bonferrroni significant MR-link-2 Mendelian randomization results, using the eQTL-Gen mRNA expression as exposures and metabolite ratios as outcomes.

**Supplementary Table 8**

Bonferrroni significant MR-link-2 Mendelian randomization results, using the UKB-PPP proteins as exposures and metabolite ratios as outcomes.

**Supplementary Table 9**

Enzyme enrichment contingency table values and Fisher exact test results per gene mapping strategy.

**Supplementary Table 10**

Metabolic pathway distance in terms of number of reactions separation (”pair distance”) between metabolite ratios (”ratio accession”). Categorized by if the metabolite ratios have an rQTL or not (”ratio has significant rqtl”). Only metabolites that are involved in an rQTL are included in the comparisons.

**Supplementary Table 11**

All metabolite ratios that exclusively contain fatty acids with a single carbon chain, and unambiguous number of double bonds. These are annotated depending on which locus they map to (*FADS*, *ELOVL* or other).

**Supplementary Table 12**

Bonferrroni and Benjamini-Yekuteli significant MR-link-2 Mendelian randomization results, using the GTEx tissue specific expression as exposures and metabolite ratios as outcomes.

**Supplementary Table 13**

Bonferrroni significant MR-link-2 Mendelian randomization results, using metabolite ratios as exposures and the finnGen complex traits as outcomes.

